# What is the recovery rate and risk of long-term consequences following a diagnosis of COVID-19? - A harmonised, global longitudinal observational study

**DOI:** 10.1101/2020.08.26.20180950

**Authors:** L. Sigfrid, M. Cevik, E. Jesudason, W.S Lim, J. Rello, J. H. Amuasi, F. Bozza, C. Palmieri, D. Munblit, J.C. Holter, A.B. Kildal, C.D. Russell, A. Ho, L. Turtle, T.M. Drake, A. Beltrame, K. Hann, I.R. Bangura, R. Fowler, S. Lakoh, C. Berry, D.J. Lowe, J. McPeake, M. Hashmi, A.M. Dyrhol-Riise, C. Donohue, D.R. Plotkin, H. Hardwick, N. Elkheir, N. Lone, A.B. Docherty, E.M. Harrison, K.J. Baille, G. Carson, M.G. Semple, J.T. Scott

**Affiliations:** Clinical Research Fellow, Public Health Specialist, ISARIC GSC, Centre for Tropical Medicine and Global Health, University of Oxford, Oxford, UK; Infection and Global Health Division, School of Medicine, University of St Andrews, St Andrews, UK; Consultant in Rehabilitation Medicine, NHS Lothian, Edinburgh, United Kingdom; Department of Respiratory Medicine, Nottingham University Hospitals NHS Trust, Nottingham, UK; Prof. 1. Centro de Investigación Biomédica en Red – Enfermedades Respiratorias (CIBERES), Vall d’Hebron Institute of Research (VHIR), Barcelona, Spain. 2. Research Department, CHU Nîmes, Université Nîmes-Montpellier, Nîmes, France; School of Public Health, Kwame Nkrumah University of Science and Technology and Kumasi Center for Collaborative Research in Tropical Medicine, Ghana; Fundação Oswaldo Cruz: Rio de Janeiro, RJ, Brasil; 1. Department of Molecular and Clinical Cancer Medicine, Institute of Systems, Molecular and Integrative Biology, University of Liverpool, UK; 2. The Clatterbridge Cancer Centre NHS Foundation Trust, Liverpool, UK; Associate Prof. 1. Department of Paediatrics and Paediatric Infectious Diseases, Institute of Child’s Health, Sechenov First Moscow State Medical University (Sechenov University), Moscow, Russia. 2. Inflammation, Repair and Development Section, National Heart and Lung Institute, Faculty of Medicine, Imperial College London, London, United Kingdom; Specialist in infectious diseases and medical microbiology, 1. Department of Microbiology, Oslo University Hospital, Oslo. 2. Institute of Clinical Medicine, University of Oslo, Oslo; Specialist in Anesthesiology, Department of Anesthesiology and Intensive Care, University Hospital of North Norway, Tromsø, Norway; Clinical lecturer in Infectious Diseases, Centre for Inflammation Research, University of Edinburgh, UK; Clinical Senior Lecturer/Consultant in Infectious Diseases, University of Glasgow, Glasgow, UK; 1. Senior clinical lecturer in infectious diseases, NIHR Health Protection Research Unit in emerging and zoonotic infections, Institute of Infection, Veterinary and Ecological Sciences, University of Liverpool. 2. Consultant in infectious diseases, Tropical and Infectious Disease Unit, Liverpool University Hospitals NHS Foundation Trust; Clinical Research Fellow, Centre for Medical Informatics, University of Edinburgh, Edinburgh, United Kingdom; Clinician, Department of Infectious Diseases, Tropical and Microbiology, IRCCS Sacro Cuore Don Calabria Hospital: Negrar di Valpolicella, Italy; Sustainable Health Systems, Freetown, Sierra Leone; Data Manager, Dorothy Springer Trust, Sierra Leone; Department of Medicine and Department of Critical Care Medicine, Sunnybrook Hospital, University of Toronto. Toronto, Canada; Prof. Institute of Cardiovascular and Medical Sciences, BHF Glasgow Cardiovascular Research Centre, University of Glasgow, Glasgow, UK; Emergency Department, Queen Elizabeth University Hospital, Glasgow, Scotland, UK; 1. NHS Greater Glasgow and Clyde 2. University of Glasgow Institute of Health and Wellbeing 3. THIS Institute, University of Cambridge; 1. Critical Care Medicine, Ziauddin University, Karachi, Pakistan 2. Pakistan Registry of Intensive CarE (PRICE), Pakistan. 3. South East Asian Research in Critical care Health (SEARCH); Prof. Dep. of Infectious Diseases, Oslo University hospital, Institute of Clinical Medicine, University of Oslo, Norway; Liverpool Clinical Trials Centre, University of Liverpool, Liverpool, UK; ISARIC Global Support Centre, Centre for Tropical Medicine and Global Health, Nuffield Department of Medicine, University of Oxford, Oxford, UK; 1. National Institute of Health Research (NIHR) Health Protection research Unit in Emerging and Zoonotic Infections, Liverpool, UK. 2. Institute of Infection and Global Health, Faculty of Health and Life Sciences, University of Liverpool, Liverpool, UK; Clinical research fellow, London School of Hygiene and Tropical Medicine, London, UK; Usher Institute, Edinburgh, UK; 1. Centre for Medical Informatics, Usher Institute, University of Edinburgh, Edinburgh, UK. 2. ^2^Intensive Care Unit, Royal Infirmary Edinburgh, Edinburgh, UK; Prof. Director Centre for Medical Informatics, Usher Institute, University of Edinburgh, Edinburgh, UK; Roslin Institute, University of Edinburgh, Edinburgh, UK; ISARIC, Centre for Tropical Medicine and Global Health, Nuffield Department of Medicine, University of Oxford, Oxford, UK; Prof. Health Protection Research Unit In Emerging and Zoonotic Infections, Institute of Infection, Veterinary and Ecological Sciences, University of Liverpool, UK 2. Respiratory Medicine, Alder Hey Children’s NHS Foundation Trust, Eaton Road, Liverpool, UK; MRC-University of Glasgow Centre for Virus Research, Glasgow, UK

**Keywords:** COVID-19, post-COVID sequelae, long-term consequences, SARS-CoV-2, observational study protocol

## Abstract

**Introduction:** Very little is known about possible clinical sequelae that may persist after resolution of the acute Coronavirus Disease 2019 (COVID-19). A recent longitudinal cohort from Italy including 143 patients recovered after hospitalisation with COVID-19 reported that 87% had at least one ongoing symptom at 60 day follow-up. Early indications suggest that patients with COVID-19 may need even more psychological support than typical ICU patients. The assessment of risk factors for longer term consequences requires a longitudinal study linked to data on pre-existing conditions and care received during the acute phase of illness.

**Methods and analysis:** This is an international open-access prospective, observational multi-site study. It will enrol patients following a diagnosis of COVID-19. Tier 1 is developed for following up patients day 28 post-discharge, additionally at 3 to 6 months intervals. This module can be used to identify sub-sets of patients experiencing specific symptomatology or syndromes for further follow up. A Tier 2 module will be developed for in-clinic, in-depth follow up. The primary aim is to characterise physical consequences in patients post-COVID-19. Secondary aim includes estimating the frequency of and risk factors for post-COVID-19 medical sequalae, psychosocial consequences and post-COVID-19 mortality. A subset of patients will have sampling to characterize longer term antibody, innate and cell-mediated immune responses to SARS-CoV-2.

**Ethics and dissemination:** This collaborative, open-access study aims to characterize the frequency of and risk factors for long-term consequences and characterise the immune response over time in patients following a diagnosis of COVID-19 and facilitate standardized and longitudinal data collection globally. The outcomes of this study will inform strategies to prevent long term consequences; inform clinical management, direct rehabilitation, and inform public health management to reduce overall morbidity and improve outcomes of COVID-19.

**Article summary:** *Strengths and limitations of this study:*

- As an international prospective, observational study we provide open-access standardised tools that can be adapted by any site interested in following up patients with COVID-19, for independent or combined analysis, to forward knowledge into short and long term consequences of COVID-19.
- This study aims to inform strategies to prevent longer term sequalae; inform clinical management, rehabilitation, and public health management strategies to reduce morbidity and improve outcomes.
- The protocol will be used for a sub-set of patients, already included in the existing cohort of more than 85,973 individuals hospitalized with confirmed COVID-19 infection across 42 countries (as of 20 July 2020), using the ISARIC/WHO standardized Core- or RAPID Case Report Forms (CRFs).
- The data will be linked with data on pre-existing comorbidities, presentation, clinical care and treatments documented in the existing cohort already documented using the ISARIC/WHO standardized Core- or RAPID CRFs.
- The data collection tool is developed to facilitate wide dissemination and uptake, by enabling patient self-assessment, however, follow up of patients requires consent and resources, which might limit the uptake and bias the data towards countries /sites with capacity to follow up patients over time.

## Introduction

Coronavirus Disease 2019 (COVID-19), caused by Severe Acute Respiratory Syndrome Coronavirus-2 (SARS-CoV-2) infection, can lead to a diverse range of clinical manifestations, ranging from an asymptomatic infection to an acute respiratory distress syndrome, and multiorgan failure with high risk of mortality (1). It is established that SARS-CoV-2 not only infects the respiratory tract but that ensuing viral replication and immune response may also affect other organs, which can lead to a risk of heart, renal and liver injury, in addition to an acute systemic inflammatory response and accompanying circulatory shock (2-4). While most people have uncomplicated recoveries, some have prolonged illness even after recovery from the acute illness (5-8). Identifying longer-term potential consequences and relationship with the acute illness is important for the management of patients, in particular, understanding how these interact and affect those already living with other conditions such as cardiovascular disease and cancer will be paramount.

However, very little is known about possible clinical sequelae that may persist after the resolution of acute infection. A recent longitudinal cohort of 143 patients followed after hospitalisation from COVID-19 in Italy, reported that 87% had at least one ongoing symptom, most (55%) with 3 or more symptoms at 60 day follow up, fatigue (53%), dyspnoea (43%), joint pain (27%) and chest pain (22%) being the most common. COVID-19 was associated with worsened quality of life among 44% of patients. (6) Prolonged course of illness has also been reported among people with mild COVID-19 who did not require hospitalisation (5, 8, 9).

Increasing evidence also suggests that infection with Sars-CoV-2 can cause neurological consequences, (2) including altered mental status, comprising encephalopathy or encephalitis and primary psychiatric diagnoses (10). While these symptoms arise acutely during the course of infection, less is known about the possible long-term consequences. Severely affected COVID-19 cases experience high levels of proinflammatory cytokines and acute respiratory dysfunction which often require assisted ventilation. These are known factors suggested to cause cognitive decline (2, 11).

Post-traumatic stress disorder (PTSD) and other consequences after intensive care unit (ICU) stay has been well documented previously (12, 13). A systematic review of consequences after hospitalisation or ICU stay for severe acute respiratory infection (SARS-CoV) and Middle East respiratory syndrome coronavirus (MERS-CoV) found consequences up to 6 months after discharge. Common consequences besides impaired diffusing capacity for carbon monoxide and reduced exercise capacity were PTSD (39%), depression (33%) and anxiety (30%) (14).

The UK government predicted in March that 45% of patients will need some form of medical or social input after recovery from COVID-19 and that 4% will require more focused, ongoing intense rehabilitation in a bedded setting (7). Early indications suggest patients with COVID-19 will need even more psychological support than typical post-ICU patients because of higher levels of “survivors’ guilt” and PTSD (15). In addition, the characteristics of the initial cellular immune and antibody response to SARS-CoV-2 have not been fully defined and it is not known if the immune responses generated by infection provides long-term protective immunity. Identifying multidisciplinary consequences through high quality, global studies throughout the course of COVID-19 is important for the acute and longer-term management of patients (4, 16).

The emerging data and anecdotal evidence of long-term recovery and persistent debilitating symptoms highlight the need for robust, standardised studies to assess the risk of and risk factors for sequalae after COVID-19. The purpose of this study is to establish a longitudinal cohort of patients with COVID-19 post-discharge, to characterize risk of long-term consequences and immune responses over time in different populations globally. This will inform strategies to prevent sequalae; inform clinical management, rehabilitation, and public health management strategies to reduce morbidity and improve outcomes.

## Methods and analysis

This protocol has been developed by the International Severe Acute Respiratory and emerging Infection Consortium (ISARIC) COVID-19 follow up working group, and informed by a wide range of global stakeholders with expertise in clinical research, outbreak research, infectious disease, epidemiology, respiratory, critical care, rehabilitation, neurology, psychology, rheumatology, cardiology, oncology and public health medicine.

### Study design

This is an international prospective, observational multi-site study to assess risk of and risk factors for longer term physical and psychosocial consequences of COVID-19 and immune response over time.

### Population and setting

This protocol builds on the ISARIC/WHO COVID-19 clinical characterization protocol and tools already in operation. The initial follow up module (Tier 1) will be used for a sub-set of patients at participating sites, already included in the existing cohort of more than 85,973 individuals hospitalized with confirmed COVID-19 infection across 42 countries (as of 20 July 2020), using the ISARIC/WHO standardized Core- or RAPID Case Report Forms (CRFs) (17). These CRFs collect data on demographics, pre-existing comorbidities and risk factors, signs and symptoms experienced during the acute phase, and care and treatments received during hospitalization. This data will be linked to the follow up data. The Core- and RAPID CRFs can be completed retrospectively for patients not yet included. New clinical sites are invited to take part in this global collaborative effort and use these open-access tools for independent or combined studies.

Specific inclusion and exclusion criteria are as follows:

### Inclusion criteria

- People aged 16 years and older
- Laboratory or physician confirmed COVID-19
- 28 days (−0/+3 months) after discharge from hospital or health centre
- Person (or family member/carer for patients who lack capacity) consent to participate

### Inclusion of vulnerable participants

The case report form and validated tools are developed for young people (>16 years of age) and adults, including pregnant women, but not validated for children. A separate case report form will be developed for children.

### Serial follow up

The follow up modules are designed in a tiered approach to be adapted depending on local resources and research needs. The Tier 1 case report form is designed to enable patient self-assessment to facilitated distribution to all patients that fit the inclusion criteria. It is developed for following up patients day 28 post-discharge, additionally at 3 to 6 months interval, with or without sampling for as many years as required (Figure 1). By being designed for patient self-completion to be administered via an online link or as a paper form, it allows wide distribution at low resource need. It can also be completed via in-clinic or telephone assessments during check-ups or for patients that are still hospitalised. The module will collect data on demographics, hospital stay and re-admissions, all-cause and cause specific mortality (after the initial index event), specific consequences including; deep vein thrombosis (DVT), pulmonary embolism, recent febrile illness, new persistent symptoms, quality of life (measured by EQ-5D-5L), dyspnoea (assessed using MRC dyspnoea scale), difficulties in functioning (UN/Washington disability score), lifestyle and socioeconomic data. The ISARIC collaborative follow up study is registered with EuroQol. Sites who wish to adopt the survey for independent studies, outside of the ISARIC collaboration need to register to use EQ-5D-5L tool with EuroQol.

**Figure 1.**
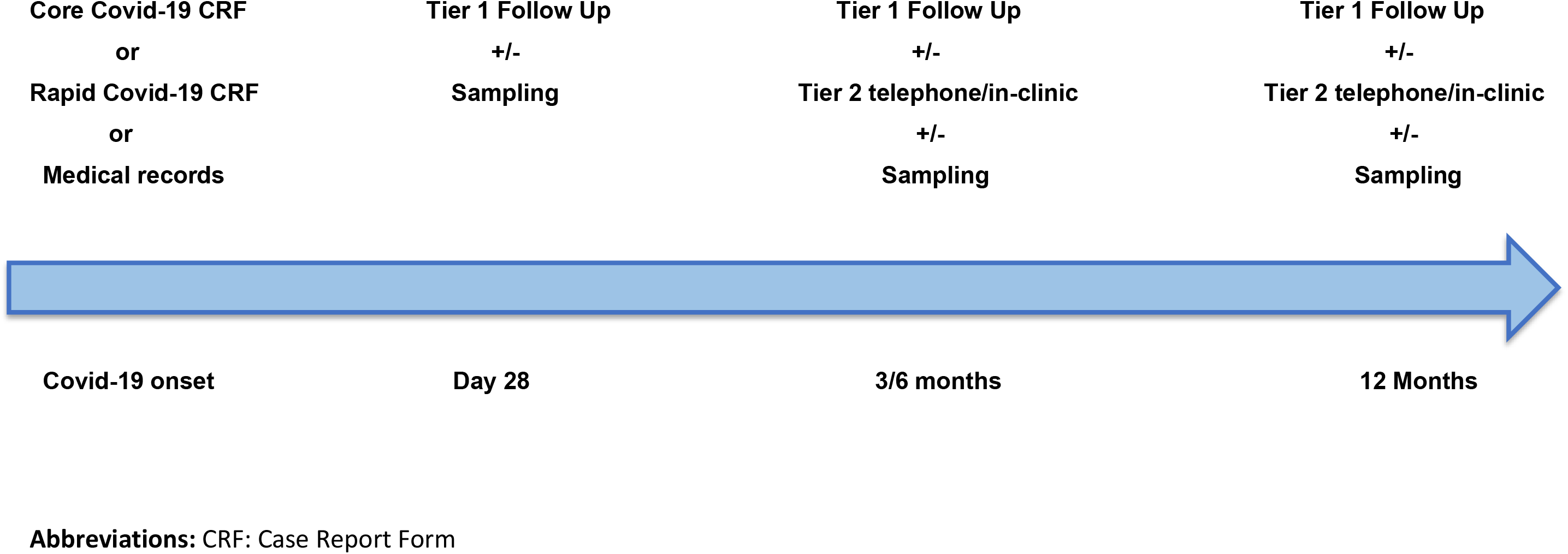
Schematic overview of the follow up data and sample time-frames.

### Sub-studies

The Tier 1 follow up module can be used to identify sub-sets of patients experiencing specific symptomatology or syndromes for further follow up. The Tier 2 follow up module will be developed for in-clinic, in-depth follow up. By using a tiered approach, additional specialist modules can be added for more complex follow up of emerging consequences in a flexible, adaptable way (Figure 2).

### Biological samples

The CRF is to be used on its own for data collection or in a sub-set of patients in combination with sampling (respiratory samples, serum, blood, stool, urine), for immunology, pathophysiology and other studies (16). It builds on the ISARIC/WHO Clinical Characterization Protocol (CCP) (18, 19). The CCP is designed for any severe or potentially severe acute infection of public health interest, such as COVID-19 (19). It is a standardized protocol for data and biological samples to be collected rapidly in a globally harmonised manner. (20) The CCP can be used for the rapid, coordinated clinical investigation of confirmed cases of COVID-19. It is designed in a tiered approach to be adapted depending on resources and includes different level of sampling schedules (acute phase and follow up) that can be adapted depending on resources, to be combined with patient data collection using the acute phase CRFs and the follow up CRF.

### Outcomes

The primary outcome of this study is to characterise physical and psychosocial consequences in patients post-COVID-19 infection. Secondary outcomes include estimating the risk of and risk factors for post-COVID-19 medical sequalae, psychosocial consequences and post-COVID-19 mortality. A subset of patients will have sampling to characterize longer term antibody, innate and cell-mediated immune responses to SARS-CoV-2 and to evaluate risk of and risk factors for re-infection with SARS-CoV-2 during re-exposure.

**Figure 2.**
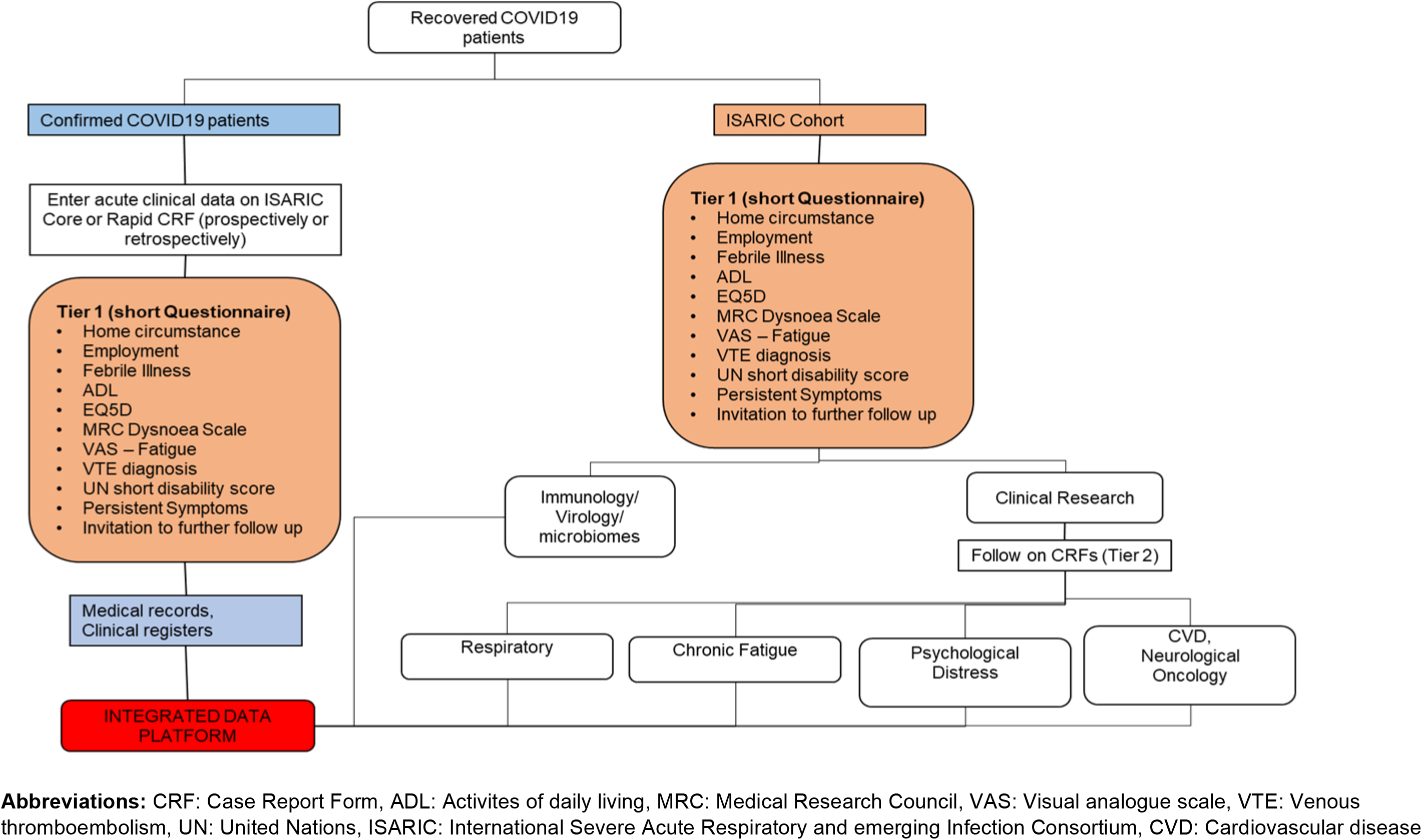
ISARIC’s adaptable Covid-19 Follow-Up Protocol framework.

### Data collection and entry

A standardized COVID-19 follow up CRF was developed through a series of virtual working group meetings and e-mail iterations. The CRF was piloted on patients in four settings in three countries and feedback incorporated into the final form (21). The CRF is designed for patient self-assessment, via online link or paper form, or to be completed during in-clinic or telephone follow-up appointment. The CRF is available open access on the ISARIC website (21) and as an electronic CRF (eCRF) on the ISARIC hosted database. The eCRF can be distributed directly to patients via an online link.

### Statistical analysis plan

Using the data, we will test for differences in outcomes across important demographic groups (age categories, sex, ethnicity, socioeconomic deprivation, comorbidities), specific exposures (severe COVID-19, critical care admission, ventilation) and initial clinical sequelae (complications on their index admission for COVID-19). We plan to use this platform to conduct timely analyses which coincide with public health or scientific need. Given these requirements, new questions we have not specified within this protocol may arise. Where this occurs, we will develop analysis plans prior to undertaking analyses, which will be made available on request. The data collected through the follow up module will be linked with data on demographics, comorbidities, clinical characteristics, care and treatments collected using the ISARIC/WHO Core- or RAPID COVID-19 CRF (15). The plan below presents our guiding statistical framework.

Entered data will be summarised first by using simple probabilistic statistics. Categorical data will be explored using frequencies and percentages, with differences in disease severity and treatment groups tested for using Chi-square tests or fisher’s exact test where cell counts are under five. For continuous data, distribution will be established using histograms and density plots. Data that are normally distributed will be summarised using group mean averages and standard deviation as a measure of central tendency. For non-parametric data, the median average will be used and presented alongside 25^th^ and 75^th^ centiles. Differences in normally distributed continuous data will be tested using Welch’s two sample t-tests for 2 group data and ANOVA for three or more groups. Mann-Whitney U will be used to compare differences across two groups or Kruskall-Wallis tests for three or more groups, where data follow a non-parametric distribution.

Outcomes will be expressed in three ways; 1) binary event data (for presence or absence of outcome of interest), 2) change over time (for continuous or ordinal data) or as 3) time to event data (for patients with serial measurements). We will calculate changes over time across symptom and outcome variables and use these changes over time to compare the effect of treatments or exposures on these outcomes. Time to event data will be captured for those who complete serial assessment forms. These data will be presented as Kaplan-Meier plots and differences tested for using log-rank tests. Competing risks (including death) will be accounted for using censoring.

To identify which patients are likely to develop persistent complications, functional impairment or reduced quality of life, we will use multilevel models to adjust for potential confounders. Level I fixed effects will include patient-level explanatory variables (i.e. age, sex) and level II or III random effects will include site and country in the case where research questions require differences in country to be accounted for. Explanatory variables will be entered into models on the basis of clinical plausibility and final model selection guided by maximisation of the adjusted R^2^ value and minimisation of the Akaike information criterion (AIC) or Bayesian information criterion (BIC). For binary event data, multilevel logistic regression will be used, and estimates presented as odds ratios alongside the corresponding 95% confidence interval. For continuous data, linear or generalised linear regression will be used, and estimates presented as model coefficients, with 95% confidence intervals. Finally, time to event data will be presented as survival probability or hazard ratios, with 95% confidence intervals.

Statistical significance will be taken at the level of P <0.05 *a-priori*. Analyses will be conducted in secure R (R Foundation for Statistical Computing, Vienna, AUT) or STATA (StataCorp LLC, TX, USA) environments.

### Data sharing

Sites who wish to utilise ISARIC’s COVID-19 data management and hosting support can to gain access to a secure data capture and management system. All systems are free to use and supported by ISARIC data management specialists. Sites who submit data to ISARIC will sign a Terms of Submission, enabling use of the data in collaborative analysis by ISARIC partners.

### Patient and public involvement

The survey has been developed with a wide range of professionals and been piloted with patients in different settings in three different countries. The feedback from the pilot has been incorporated into the final version of the Tier 1 survey. Since follow-up modules will involve both phone/online methods and will also include a subset who will come in-person, we aim to consult local community-based groups and organizations on the best approaches to achieving high levels of participation and limiting the attrition rate.

## Ethics and dissemination

Since the emergence of SARS-CoV-2 in December 2019, there has been a considerable global effort to characterise the virus and clinical course of disease. This has included identification of the virus, rapid design and development of diagnostics, host cell receptor identification, and insights into early epidemiological and clinical parameters. However, very little is known about possible clinical sequelae that may persist after resolution of the acute COVID-19. Emerging data indicates impact of COVID-19 infection on not only the respiratory system but also, the kidneys, liver, neurological, psychological and multisystem inflammatory syndrome (2, 4, 10, 22). Moreover, with increasing reports of long-term consequences, there is an urgent need to characterise the risk of short and chronic consequences and risk factors and biomarkers for patients at risk of sequelae, to inform preventative and rehabilitation strategies.

The assessment of risk factors for longer term consequences requires a longitudinal study linked to data on pre-existing conditions and patient data and care received during the acute phase. A subset of patients included in the ISARIC/WHO clinical characterisation protocol has provided consent to be contacted for follow up. Additionally, sites taking part in the global follow up study will apply for consent to follow up patients confirmed with COVID-19, following the local ethical committee procedures. These standardised protocols and tools will serve as a standardised template that can be modified as appropriate and needed at each site. With this study we aim to characterize the risk of long-term consequences and nature of innate and adaptive immune responses over time in patients following a diagnosis of COVID-19. We will collect data on wide range of outcomes including hospital stay and re-admissions, all-cause and cause specific mortality (after the initial index event), consequences e.q. DVT, pulmonary embolism, recent febrile illness, new, persistent symptoms, EQ-5D-5L, MRC dyspnoea scale, UN/Washington disability score, lifestyle and employment data. Data from combined analysis will be disseminated through the ISARIC website and in open-access publications, under group authorship.

The follow up module is developed as an open-access, flexible tool to be adopted and adapted as appropriate depending on need and resources by any site globally interested in following up patients with COVID-19 over time, to facilitate standardized data collection globally and combined analysis. The outcomes of this study will inform strategies to prevent risk of consequences; clinical management, rehabilitation, and public health management, to reduce morbidity and improve outcomes. By standardising data collection, and providing open-access, adaptable tools, focused on key data variables, it can optimize data quality and reduce the burden of research on staff, while collecting the most relevant information to inform clinical care guidelines and public health. The tools will be made available in a range of languages.

### Data statement

ISARIC stands by the principles of data sharing in public health emergencies. ISARIC supported studies will share quality data in a timely, valid, and governed manner to inform public health policy and benefit patient care. The ISARIC-hosted data platform enables rapid and harmonised operationalisation of data collection to a secure database (23). Ownership and control of the data entered are retained by those who enter the data. Sites can contribute data to combined analysis upon permission. Technical appendix, statistical code and datasets are available from https://isaric.tghn.org/ or by contacting. Research outcomes will be disseminated via the ISARIC website and published in peer-reviewed scientific journals, under a collaborative group authorship. The data contributors will share the outcome results with key stakeholders to inform public health response, policy development and implementation. To efficiently make findings from this project visible and accessible to a wide range of stakeholders as well as to networks of individuals and institutions, we will utilise social media, including the ISARIC and partner institution’s websites. The consortium will manage a page dedicated to this project, that will incorporate all the tools available. The webpage will provide not only updates on the progress of the project and component activities and events, but also information on research.

This follow up study is developed as an open-access tool to be adopted as appropriate by any site interested in following up patients with COVID-19 over time, to facilitate standardized data collection globally to enable combined analysis. New sites globally are invited to join the study at any time. The outcomes of this study will inform strategies to prevent risk of consequences; clinical management, rehabilitation and public health management needs to reduce morbidity and improve outcomes.

## Data Availability

ISARIC stands by the principles of data sharing in public health emergencies. ISARIC supported studies will share quality data in a timely, valid, and governed manner to inform public health policy and benefit patient care. The ISARIC-hosted data platform enables rapid and harmonised operationalisation of data collection to a secure database. Ownership and control of the data entered are retained by those who enter the data. Sites can contribute data to combined analysis upon permission. Technical appendix, statistical code and datasets are available from https://isaric.tghn.org/ or by contacting.

https://isaric.tghn.org/CCP/isaric-global-covid-19-long-term-follow-study/

## Acknowledgments

We would like to thank the ISARIC global clinical characterisation group, ISARIC4C and all the clinicians, nurses and researchers contributing COVID-19 clinical patient data and samples, which will be linked with the follow up data and all the patients that have consented to be followed up. We would like to acknowledge WHO, whose working groups contributed to development of the CCP and associated tools. Moreover, Sarah Moore, Romans Matulevics, Katherine Maskell, Peter Bannister, Liliana Resende and Anneli Sandström for administrative, graphic design and dissemination support.

## Author contributions

JTS and LS lead on the development of the follow up protocol and tools in collaboration with members of the ISARIC COVID-19 follow up working group (JR, JHA, FB, CP, DM, JCH, ABK, A-MDR, KH, IRB, AB, MH, RF, SL, AH, HH, CD, NE, LT, CR, MC) and external specialists (EJ, WSL, DJL, CB, JM, NL). TD, LS, JTS, ABD, EH developed the statistical analysis plan. DP managed the database set up. AH, LT, ABK, JR, CP piloted the CRF. MGS, KJB, EH, ABD, GC provided scientific advice. LS and MC lead on drafting of the protocol and manuscript, with contributions from all co-authors. All authors reviewed and approved the final manuscript.

## Competing interest statement

No competing interests declared.

## Funding statement

This work was supported by the Department for International Development and Wellcome [215091/Z/18/Z] and the Bill & Melinda Gates Foundation [OPP1209135]. CP would like to acknowledge the support of the Liverpool Experimental Cancer Medicine Centre (Grant Reference: C18616/A25153) and The Clatterbridge Cancer Centre Charity. CB acknowledges the support from British Heart Foundation RE/18/6134217.

## Notes

### Competing Interest Statement

The authors have declared no competing interest.

### Clinical Trial

osf.io/c5rw3/

### Clinical Protocols

https://isaric.tghn.org/CCP/isaric-global-covid-19-long-term-follow-study/

### Author Declarations

This is a multi-site global prospective COVID-19 observational follow up study protocol, developed by the ISARIC COVID-19 Global Follow up working group. Each site that sets up the study will seek local ethical approvals as appropriate.

